# Possibility of SARS-CoV-2 infection in metastatic microenvironment of cancer

**DOI:** 10.1101/2021.05.24.21257662

**Authors:** Takuma Hayashi, Ikuo Konishi

## Abstract

According to a report from the World Health Organization, the mortality and severity rates among patients with cancer infected with severe acute respiratory syndrome coronavirus 2 (SARS-CoV-2) are significantly higher than those of individuals infected with SARS-CoV-2 without complications. Common and cancer-specific risk factors may be involved in the mortality and severity rates of coronavirus disease 2019 (COVID-19). Various factors have been determined to contribute to the aggravation of COVID-19 in patients with cancer. However, on the basis of current research, the factors involved in the aggravation of COVID-19 in patients with cancer have not been fully investigated. In the general course of treatment for patients with cancer, the detection of the formation of metastases in other organs is common. Therefore, the present study investigates the association between lung metastatic lesion formation and SARS-CoV-2 infectivity. On the basis of the results obtained, in the pulmonary micrometastatic niche of patients with ovarian cancer, alveolar epithelial stem-like cells adjacent to the ovarian cancer were observed. Moreover, it was revealed that angiotensin-converting enzyme 2, a host-side receptor for SARS-CoV-2, was expressed in alveolar epithelial stem-like cells adjacent to the ovarian cancer in the pulmonary micrometastatic niche. Furthermore, it was also observed that the SARS-CoV-2 spike glycoprotein receptor-binding domain binds to alveolar epithelial stem-like cells. In other words, it was suggested that patients with cancer and pulmonary micrometastases may be more susceptible to SARS-CoV-2. The prevention of *de novo* niche formation in metastatic disease may be a new strategy for the clinical treatment of COVID-19 for patients with cancer.

## Introduction

The United States and other countries have found it difficult to contain the coronavirus disease 2019 (COVID-19) pandemic owing to the respiratory spread of severe acute respiratory syndrome coronavirus 2 (SARS-CoV-2) and the inconsistent adherence to effective public health measures, including wearing masks and maintaining social distancing. According to the reports of the World Health Organization, the mortality rate of patients with cancer infected with SARS-CoV-2 is 7.6%, which is a fairly high rate compared with the 1.4% mortality rate of SARS-CoV-2–infected individuals without complications^1^. Among patients with cancer and COVID-19, the 30-day all-cause mortality was high and associated with general and cancer-specific risk factors with a mortality rate of 13.3%^1,2^. Moreover, the severity rate of patients with solid cancer infected with SARS-CoV-2 as of August 2020 was extremely higher than that of all the SARS-CoV-2–infected patients according to a report by the Japan Ministry of Health, Labour and Welfare^3^. The reason why COVID-19 becomes more severe in patients with cancer is not fully understood. Consequently, immunity against the virus may be reduced in patients with cancer receiving therapeutic anticancer agents^4^.

Lung stem cells that can regenerate lung tissue are used in research on lung disease, cancer, and infectious diseases. Furthermore, preparing a lung culture model using lung stem cells in the laboratory has become possible. However, the biological characteristics of human lung stem cells have not been clarified. Therefore, research on the construction of a lung culture model, especially the model of the most peripheral part of the lung where gas exchange is observed, has not progressed.

Research findings on human lung culture systems have been reported to model lung infections, including SARS-CoV-2 infections that are responsible for COVID-19-associated pneumonia^5,6^. Moreover, Kuo et al. succeeded in creating a human peripheral lung culture model using lung stem cells^5^. Epithelial cell adhesion molecule-positive alveolar epithelial type 2 (AT2) stem cells differentiate into AT2 progenitor cells expressing angiotensin-converting enzyme 2 (ACE2), which plays a role as a receptor for SARS-CoV-2. In contrast, cytokeratin 5-positive lung stem cells differentiate into bronchial and/or bronchiolar epithelial progenitor cells. In other words, the AT2 progenitor cells in a human peripheral lung culture model express ACE2, a host cell receptor for SARS-CoV-2^5^. Thus, SARS-CoV-2 infects alveolar epithelial stem cells.

In this study, it was shown that ACE2 was expressed in alveolar epithelial stem-like cells adjacent to the ovarian cancer in the pulmonary micrometastatic niche. Furthermore, it was also observed that the receptor-binding domain (RBD) of spike glycoprotein of SARS-CoV-2 binds to alveolar epithelial stem-like cells. In other words, indicating that patients with cancer and pulmonary micrometastases might be more susceptible to SARS-CoV-2. Prevention of *de novo* niche formation in metastatic disease may be a new strategy for the clinical treatment of COVID-19 for patients with cancer.

## Materials and Methods

### 1. Case selection for immunohistochemical staining

To examine the biological and medical characteristics of pulmonary micrometastatic niche, four cases (high-grade serous ovarian adenocarcinomas) were selected from a total of 69 primary epithelial ovarian cancers stained via immunohistochemical analysis with anti-human S100 calcium-binding protein A4 (S100A4), which is used as a marker for ovarian cancer (Supplementary Table 1). Sixty-nine consecutive patients with ovarian carcinoma visited Shinshu University Hospital (Matsumoto, Nagano, Japan) between 1994 and 2003 and underwent surgery followed by combination chemotherapy with taxane-based preparation and platinum preparation. The supplementary materials show the detailed medical conditions of the patients.

### 2. Antibodies and immunohistochemistry (IHC)

IHC staining for S100A4, CD90(Thy1), ACE2, and RBD of the spike glycoprotein of SARS-CoV-2 was performed on the tissue sections of pulmonary micrometastases of patients with high-grade serous ovarian cancer. Tumor tissue sections were then incubated with the appropriate primary antibodies at 4 °C overnight. These IHC experiments with human tissue sections were conducted using standard procedures at Shinshu University (Matsumoto, Nagano, Japan) and National Hospital Organization Kyoto Medical Center (Kyoto, Kyoto Japan) in accordance with institutional guidelines (approval no. M192). The supplementary materials show the list of antibodies, which were used as the primary or secondary antibody in our research experiments, as well as the detailed materials and methods.

## Results

Niches for promoting metastatic colonization were previously examined using the generation of human-in-mouse ovarian cancer xenograft models in immunodeficient mice; i.e., CD34^+^ lineage ovarian cancer stem-like cells sorted using the side population procedure were injected into the mammary fat pads of BALB/c *nu/nu* mice. The cluster of differentiation 90 (CD90), which is also called Thy1, is used as a marker for several stem cells^7^. S100 calcium-binding protein A4 (S100A4), a member of the S100 calcium-binding protein family secreted by ovarian cancer cells, supports tumorigenesis by stimulating angiogenesis^7^ (Supplementary Table 1). Pathological examinations revealed the existence of S100A4-negative and CD90-positive stem-like cells in vimentin-positive normal neighboring alveolar epithelial cells^8^. In the present study, our experiments demonstrate that similarly, the initialization of mimicry represented the incomplete differentiation of normal alveolar epithelial cells toward the stem-like lineage in the pulmonary micrometastasis of patients with ovarian cancer (Figure 1).

**Figure 1.**
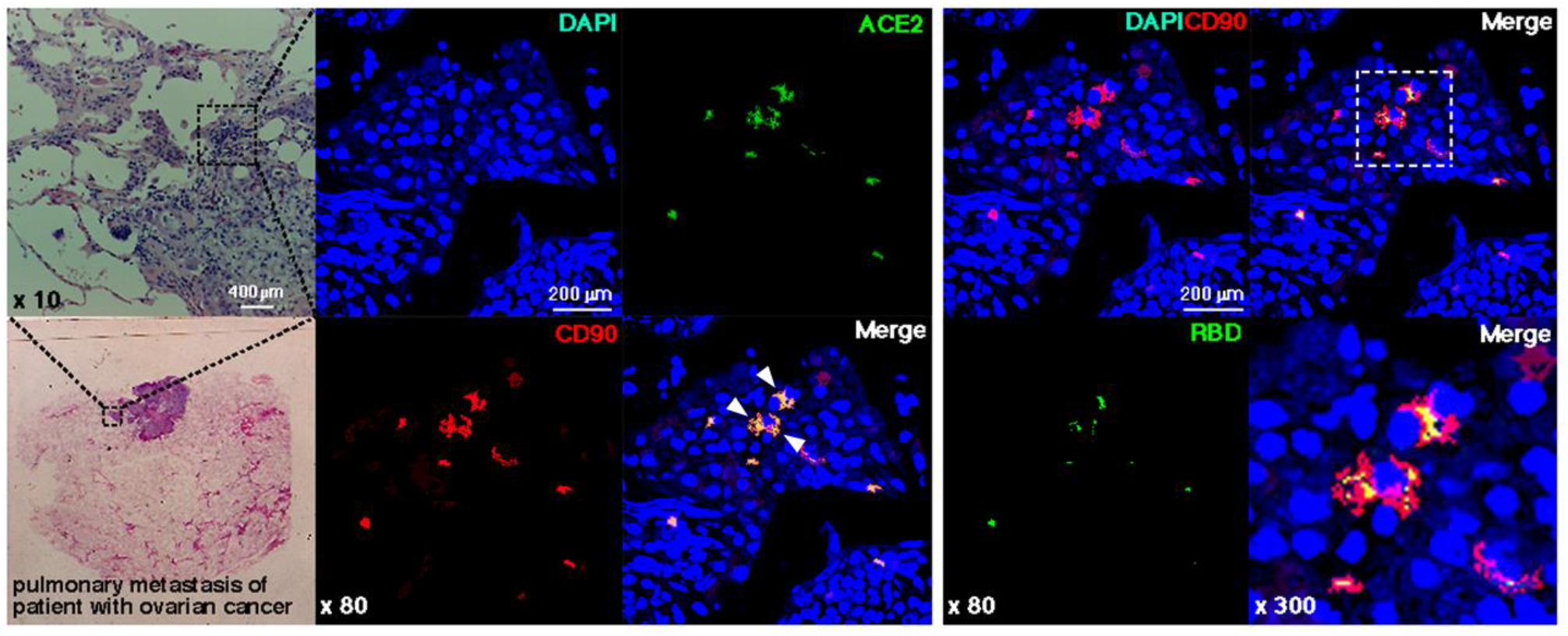
Binding of RBD in the spike glycoprotein of SARS-CoV-2 to stem-like cells in normal neighboring alveolar epithelial cells. Immunohistochemical studies were performed using pulmonary metastasis tissue surgically excised from a patient with serous ovarian carcinoma. The existence of human ACE2-positive (*green*) and human CD90-positive (*red*) stem-like cells indicated by *white arrowheads* in human normal neighboring alveolar epithelial cells was observed in pulmonary micrometastasis. Human CD90-positive cells (*red*) are not detected in the metastatic colonies of human serous ovarian carcinoma. Immunohistochemical studies were performed using an antibody for human ACE2 (*green*), a monoclonal antibody for the spike glycoprotein of SARS-CoV-2 (*green*), and an antibody for human CD90 (*red*) as a biomarker for stem-like cells. The binding of RBD in the spike glycoprotein of SARS-CoV-2 (*green*) to CD90-positive (*red*) alveolar epithelial stem-like cells is observed, which are indicated in *yellow*. Antihuman CD90 antibody (Abcam ab133350), antihuman ACE2 antibody (ORIGENE, Rockville, MD, USA), anti-spike glycoprotein of SARS-CoV-2 antibody (GeneTex, Inc., CA, USA), and recombinant spike glycoprotein of SARS-CoV-2 protein (BioVision, CA, USA) are used in immunohistochemistry studies. The experiments were performed five times with similar results.

The expression of ACE2, a host-side receptor for SARS-CoV-2, in CD90-positive alveolar epithelial stem-like cells in the pulmonary metastatic niches of patients with high-grade serous ovarian cancer is essential (Figure 1 and Table 1). Furthermore, histopathological analysis revealed that the RBD in the spike glycoprotein of SARS-CoV-2 binds to ACE2-expressing CD90-positive alveolar epithelial stem-like cells (Figure 1 and Table 1). Moreover, on the basis of such findings, SARS-CoV-2 is deemed to infect the alveolar epithelial stem-like cells in the pulmonary micrometastasis of patients with ovarian cancer.

**Table 1.**
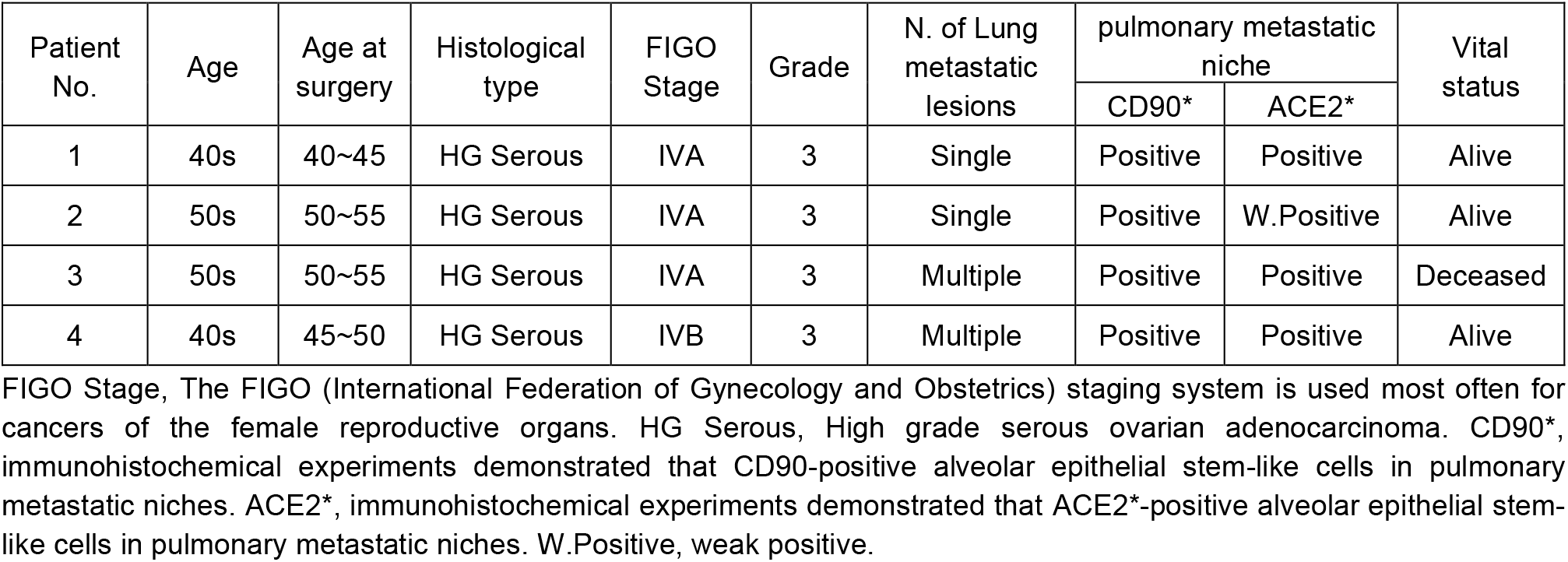
Patient characteristics ovarian cancer with lung metastasis and CD90, ACE2 expression

From the results of previous studies, it has been reported that in the pulmonary micrometastatic niche of other cancer types, the reprogramming of mimicry probably represents incomplete differentiation of normal alveolar epithelial cells toward stem-like lineages. Presumably, pulmonary micrometastatic niche is a target for SARS-CoV-2 infection.

## Discussion

The results showing that ovarian cancer cells form a metastatic niche near the alveolar stem cells is reminiscent of a previous finding that demonstrates that when prostate cancer cells metastasize to the bone, they settle near the stem cells in the bone marrow, which promotes the development of a metastatic environment that supports tumor growth^9^. A recent report also described cancer-associated parenchymal cells that exhibit stem cell-like features, the expression of lung progenitor markers, multilineage differentiation potential, and self-renewal activity^10^.

Among patients with cancer and COVID-19, 30-day all-cause mortality has been reported to be high, and various factors have been determined to contribute to COVID-19 severity in patients with cancer. Within the pulmonary metastatic niche, alveolar epithelial cells adjacent to metastatic cancer cells are initiated into alveolar epithelial stem-like cells. ACE2 has been expressed in alveolar epithelial stem-like cells, and the binding of alveolar epithelial stem-like cells to RBD of the spike glycoprotein of SARS-CoV-2 is clearly observed. One of these factors is considered to be SARS-CoV-2 infection of stem cells and/or epithelial progenitor cells present in the metastatic niche in patients with cancer. So far, treatment aimed at reducing lung metastases has been limited to surgical treatment. However, in clinical practice, the reduction of lung metastases has been observed in the treatment with immune checkpoint inhibitors and/or PARP inhibitors. Furthermore, the efficacy of the anti-S100A4 antibody drug in suppressing the metastatic ability of malignant tumors in other organs, including ovarian cancer, has been investigated^11^. A longer follow-up is needed to better understand the effect of COVID-19 on the treatment outcomes of patients with cancer, including the ability to continue specific cancer treatments. In such a major intersection of cancer medicine and infectious diseases, the prevention of *de novo* niche formation of metastatic disease may be a novel strategy for the clinical treatment of COVID-19.

## Data Availability

The materials and methods used in this study are listed in the supplementary material.

## Author contributions

TH performed most of the clinical treatments and diagnostic pathological studies, coordinated the project, created the study, and wrote the manuscript. IK carefully reviewed the manuscript and commented on the aspects of clinical medicine. IK shared information on clinical medicine and oversaw the entirety of the study.

## Footnote

The material (manuscript and figure) is original research. It has not been previously published and has not been submitted for publication elsewhere while under consideration.

## Data availability and Consent to publish

This manuscript is an editorial and does not contain research data.

Therefore, there is no research data or information to be published or opened.

## Ethical approval and consent to participate

This research study (Research number: UMIN000038789) was reviewed and approved by the Central Ethics Review Board of the National Hospital Organization Headquarters in Japan (Tokyo Japan). The authors attended a 2020 educational lecture on medical ethics supervised by the Japanese government. The completion numbers of the authors are AP0000151756 AP0000151757 AP0000151769 AP000351128. This research is clinical research (Research number: UMIN000038789), therefore consent to participate is required, therefore we have received a written consent from the subjects who participated in the clinical research.

## Research funding

This clinical research was performed with the support of the following research funding: Japan Society for the Promotion of Science for TH (grant No. 19K09840), START-program Japan Science and Technology Agency (JST) for TH (grant No. STSC20001), and National Hospital Organization Multicenter clinical study for TH (grant No. 2019-Cancer in general-02).

## Acknowledgments

We thank all the medical staffs and co-medical staffs for their contribution to the medical research at National Hospital Organization Kyoto Medical Center. We appreciate Crimson Interactive Japan Co., Ltd., for revising and polishing our manuscript.

