## supplemental materials for "Possibility of SARS-CoV-2 infection in metastatic microenvironment of cancer"

Takuma Hayashi, Kenji Sano, Nobuo Yaegashi, Ikuo Konishi

### Materials and Methods

#### *1. Antibodies*

List of antibodies, which were used as first monoclonal antibody or secondary antibody in our immunohistochemistry research experiments, is shown below.

**Antibodies**

| Antibody | Company | Catalogue No. | Clonal (Clone) | Specificity | Dilution |
|---|---|---|---|---|---|
| S100A4 | Abcam | ab124805 | Rabbit monoclonal EPR2761(2) | Human | 1:200 (IHC) |
| CD90 | Abcam | ab133350 | Rabbit monoclonal EPR3133 | Human | 1:200 (IHC) |
| ACE2 | ORIGENE | CF803844 | Mouse monoclonal 4C5 | Human | 1:150 (IHC) |
| RBD of Spike | GeneTex | GNT-9366-04 | Mouse monoclonal 1A9 (GNT936604) | SARS-CoV-2 | 1:200 (IHC) |
| Anti-mouse IgG Alexa Fluor® 488 | Invitrogen | A32723 | Goat / IgG | Mouse IgG | 1:100 |
| Anti-Rabbit IgG Alexa Fluor® 546 | Invitrogen | A-11035 | Goat / IgG | Rabbit IgG | 1:200 |

#### *2. Case selection for immunohistochemical staining*

Cases were selected from total of 69 primary epithelial ovarian cancers for immunohistochemical analysis. Sixty-nine consecutive patients with ovarian carcinoma visited Shinshu University Hospital between 1994 and 2003 and underwent surgery followed by cisplatin-based chemotherapy. The follow-up period ranged from 3 to 160 months (median: 76 months). According to the International Federation of Gynecology and Obstetrics (FIGO) classification, 33 carcinomas were classified as Stage I, 10 were classified as Stage II, 21 were classified as Stage III and 5 were classified as Stage IV. Ovarian epithelial cancers are classified into serous, mucinous, endometrioid, clear cell, transitional cell, squamous cell, mixed epithelial and undifferentiated categories depending on histomorphologic features[1]. Histologically, 25 were serous, 5 were mucinous, 22 were clear cell and 17 were endometrioid adenocarcinomas (Supplementary Table 1). In 24 of the 69 cases, the specimens of peritoneal dissemination were available and also examined for immunohistochemistry. The expression of S100A4 in the nucleus and cytoplasm in all 69 cases was examined by immunohistochemical staining (Supplementary Table 1). Serous ovarian cancer has a lower 5-year survival rate than other histological types (Supplementary Table 1). Of the five cases at stage IV, two cases of serous ovarian carcinoma were available for lung metastases and were also examined for immunohistochemistry. Each tissue sample was used with the approval of the Ethics Committee of Shinshu University School of Medicine.

***3. Immunohistochemistry (IHC)***

IHC staining for CD90(Thy1), S100A4, ACE2, and RBD of spike glycoprotein of SARS-CoV-2 was performed on tissue sections of ovarian cancers. Antibodies for CD90(Thy1) (ab133350), S100A4 (ab124805) were purchased from Abcam Inc. (Cambridge, UK). RBD of spike glycoprotein was purchased from GeneTex Inc. (Irvine CA USA). DAPI Mounting Medium was purchased from VECTOR LABORATORIES, Inc. (Burlingame, CA). IHC was performed using normal methods with the primary antibody and second antibody conjugated with immunofluorescence as described previously. Briefly, one representative 5-µm-thick tissue section was cut from a paraffin-embedded sample derived from patients with ovarian cancer. To examine the expression levels of target molecules, we performed immunofluorescence experiments for CD90(Thy1), S100A4 on paraffin-embedded xenografts derived from patients with ovarian cancer. Tumor tissue sections were then incubated with the appropriate primary antibodies at 4°C overnight. We used rabbit monoclonal antibodies to S100A4 (1:200), a rabbit monoclonal antibody to CD90(Thy1) (1:200), a mouse monoclonal antibody to human ACE2 (1:150), and RBD of spike glycoprotein of SARS-CoV-2 (1:200) as the primary antibody. After being incubated with the secondary antibody, i.e., the Alexa Fluor® 488-conjugated anti-mouse IgG antibody or Alexa Fluor® 546-conjugated anti-rabbit IgG antibody (1:200; Invitrogen), sections were washed and cover slipped with mounting medium and 40,6-diamidino-2-phenylindole (DAPI) (Vectashield; Vector Laboratories) and then visualized under a confocal microscope (Leica TCS SP8, Wetzlar, Germany) according to the manufacturer's procedure. Normal rabbit or mouse antiserum was used as a negative control for the primary antibody. These experiments with human tumor tissues derived from patients with ovarian cancer were conducted at Shinshu University and National Hospital Organization Kyoto Medical Center in accordance with institutional guidelines (approval no. M192).

1. https://www.proteinatlas.org/learn/dictionary/pathology/ovarian+cancer

**Supplementary Table 1**

| Histological type | Total cases | Expression of S100A4 | | | | | | 5-year survival rate |
|---|---|---|---|---|---|---|---|---|
| | | Cytoplasmic staining | | | Nuclear staining | | | |
| | | + | ++ | +++ | + | ++ | +++ | |
| Serous | 25 | 0 | 3 | 22 | 11 | 6 | 8 | 59.9% |
| Mucinous | 5 | 0 | 1 | 4 | 1 | 3 | 1 | 91.1% |
| Endometrioid | 17 | 3 | 4 | 10 | 9 | 4 | 4 | 81.3% |
| Clear cell | 22 | 2 | 5 | 15 | 13 | 5 | 4 | 77.5% |

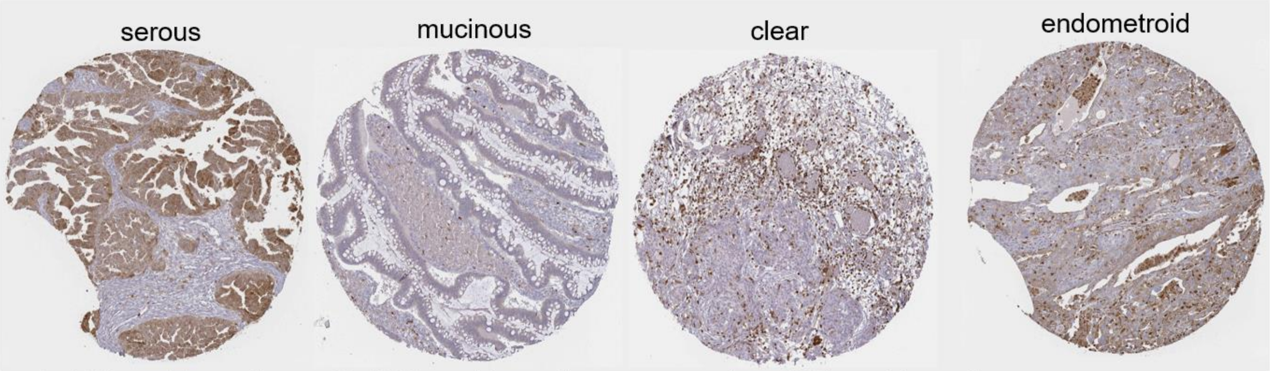


–, 0–10% positive cells; +, 10–50% positive cells; + +, more than 50% positive cells.
